# Genetic and atherosclerotic plaque immunohistochemical analyses do not associate reduced sclerostin expression with cardiovascular events

**DOI:** 10.1101/2020.11.20.20235234

**Authors:** Gill Holdsworth, James R Staley, Peter Hall, Ian van Koeverden, Ciara Vangjeli, Remi Okoye, Rogely Boyce, James R Turk, Martin Armstrong, Alison Wolfreys, Gerard Pasterkamp

## Abstract

The sclerostin antibody romosozumab increases bone formation and decreases bone resorption, leading to increased bone mass, bone mineral density and bone strength, and reduced fracture risk. In a clinical study versus alendronate in postmenopausal women (ARCH), an imbalance in adjudicated serious cardiovascular (CV) events driven by an increase in myocardial infarction (MI) and stroke was observed.

To investigate whether inhibition of sclerostin in atherosclerotic plaques may have contributed to this imbalance, sclerostin was immunostained in human plaques to determine whether it was detected in regions relevant to plaque stability. Additionally, genetic variants associated with lifelong reduced sclerostin expression were explored for associations with phenotypes including those related to bone physiology and CV risk factors/events in a population-based phenome-wide association study (PheWAS).

Sclerostin expression was absent (67%) or reduced in atherosclerotic plaques and when present was in deeper regions of the plaque/wall and not in areas considered relevant to plaque stability (fibrous cap and endothelium). Natural genetic modulation of sclerostin by variants with a significant positive effect on bone physiology showed no association with lifetime risk of MI or stroke. These data do not support a causal association between sclerostin inhibition and increased risk of serious cardiovascular events.

## 1. Introduction

Sclerostin is the secreted protein product of the *SOST* gene, which inhibits canonical Wnt signalling in the skeleton, acting as a negative regulator of bone formation (1). Genetic deficiency or absence of sclerostin causes the rare high bone mass conditions sclerosteosis and van Buchem disease (2, 3). In adults, sclerostin is principally expressed by skeletal osteocytes (4). Antibodies to sclerostin, such as romosozumab, increase bone formation and decrease bone resorption, leading to increased bone mass, bone mineral density (BMD) and bone strength, and reductions in fracture risk (5).

*SOST* mRNA/sclerostin protein are also constitutively expressed in aorta (3, 6-8) and upregulated in foci of vascular calcification (9-14). The function of sclerostin in the vasculature is unclear, however it has been previously been proposed as a potential inhibitor of vascular calcification (9, 14). Since excess sclerostin has been reported to be protective against atheroprogression and inflammation in an ApoE mouse model (8), it has also been hypothesised that a reduction in sclerostin could be associated with an increased risk of atheroprogression. However, a recently published study found no association between sclerostin inhibition and atheroprogression when reduction in sclerostin was directly assessed in two ApoE mouse models using different methods of inducing atheroprogression (15). In the recent ARCH phase III comparator clinical trial of romosozumab or alendronate in women with post-menopausal osteoporosis, a numerical imbalance in positively adjudicated serious cardiovascular adverse events was observed during the first 12 months of the study (50 patients (2.5%) in romosozumab group vs. 38 patients (1.9%) in the control alendronate group) (16), which was driven by a greater number of myocardial infarction and stroke events. This imbalance was not observed in the larger placebo-controlled FRAME phase III clinical trial (17). A comprehensive nonclinical toxicology package with additional cardiovascular studies has been conducted on romosozumab (15) and whilst the expected pharmacodynamic effects were observed in bone, there were no functional, morphological, or transcriptional effects on the cardiovascular system in animal models in the presence or absence of atherosclerosis. At present, a biologically plausible mechanistic link between romosozumab and an imbalance of CV events (MI and stroke) has not been identified.

Atherosclerosis is a chronic progressive inflammatory disease of the arterial wall that is a significant cause of morbidity and mortality (18). Sudden changes in advanced atherosclerotic (AS) plaques, such as plaque erosion or rupture with thrombosis, or arterial spasm in the presence or absence of plaque (19, 20) are common pathological mechanisms associated with adverse cardiovascular events such as myocardial infarction and stroke. Plaque rupture is usually associated with the ‘vulnerable’ plaque morphology (21). However, superficial erosion of fibrous plaques, which is more frequently observed in women, is recognised as a significant driver of acute coronary deaths (22-24).

Little is known about sclerostin expression in AS plaques. Here, immunohistochemistry (IHC) was used to assess sclerostin expression in advanced AS plaques collected from older female patients and stored in the AtheroExpress biobank (25). This enabled assessment of sclerostin expression in the atherosclerotic plaques in women of a similar age range to that of the romosozumab phase III clinical trials (ARCH and FRAME), and investigation into whether sclerostin staining was associated with histological features of plaque instability, intra-plaque cytokine profile or patient demographics: age at endarterectomy, history of arterial disease pre-surgery, or cardiac events during a three-year follow-up period.

To further explore any mechanistic links between reduced sclerostin and cardiovascular outcomes, population-based genetic approaches were employed. Phenome-Wide Association Studies (PheWAS) allow unbiased, systematic assessment of causative associations between common human genetic variants and multiple human diseases and phenotypic traits (26). Although the effect sizes of complex trait genetic associations tend to be small in magnitude (27) (especially when compared to drug effects), this approach allows naturally occurring common genetic variation within a population to be used as a surrogate for the action of a drug, providing supporting data to predict both efficacy and on-target safety profiles (28, 29). Here, variants in the *SOST* region associated with reduced expression of the *SOST* gene, encoding sclerostin, and increased BMD were selected as a proxy for sclerostin inhibition and used in PheWAS to assess the phenotypic associations for these variants at a population level. Implicitly, if the variants reflect the pharmacological action of sclerostin inhibition then associations with increased BMD and reduced risk of fractures would be expected. Therefore, the same variants can be used to explore any mechanistic link between genetically reduced sclerostin levels and cardiovascular outcomes.

## 2. Results

### 2.1. Immunohistochemistry of atherosclerotic plaques

#### 2.1.1. Plaque Characteristics

We selected a random sample of 94 carotid and 50 femoral/iliac artery plaques acquired from female patients of an age which meant they were likely to be post-menopausal (mean age 69.6 years ± 10.4), and archived as formalin-fixed paraffin-embedded (FFPE) blocks in the AtheroExpress biobank. The characteristics of the study group are summarised in Table S1.

Plaques had been collected during surgical endarterectomy from patients showing 50-95% stenosis. In this procedure, the plaque and adjacent tunica intima (T. intima) was dissected away leaving a significant proportion of samples with residual tunica media (T. media), confirmed by hematoxylin and eosin (H&E) and alpha smooth muscle actin (αSMA) staining. T. media was present in 97% of carotid artery plaques and 78% of femoral/iliac samples. Plaques were classified as previously described (25): carotid plaques were categorized as atheromatous (33%), fibroatheromatous (16%) or fibrous (51%) whilst femoral/iliac plaques were typically fibrous (88%) with the remainder being fibroatheromatous.

#### 2.1.2. Sclerostin antibody optimisation and specificity

Careful optimisation of the sclerostin IHC protocol was critical to ensure the specificity of staining and hence the reliability and reproducibility of this study. Two proprietary sclerostin antibodies (Scl-Ab) and two commercially available Scl-Ab were evaluated for their ability to sensitively and specifically stain sclerostin protein in FFPE tissue sections. Human aorta was used as a positive control tissue and human kidney and liver as negative control tissues. Kidney expresses *SOST* mRNA (2, 3) but not sclerostin, whilst *SOST* is not detected in liver (2).

All four Scl-Ab tested showed strong staining of vascular smooth muscle cells (VSMC) in aorta T. media, however three of the four antibodies exhibited non-specific (background) staining in the negative control kidney and liver sections, where sclerostin is not expressed (Figure S1), thus their use would generate misleading data on tissue protein expression. Only one antibody (Scl-Ab #1) showed specific and sensitive staining of positive control tissues and absent staining of the negative control section, and hence was suitable for IHC applications. This antibody was used to stain the 144 AS plaques selected from the AtheroExpress Biobank.

#### 2.1.3. Sclerostin expression in normal aorta and atherosclerotic plaques from the AtheroExpress Biobank

Using an optimised sclerostin staining protocol for the selected antibody, intense sclerostin immunoreactivity was observed in two control FFPE normal aorta specimens, whilst an isotype negative control antibody showed no staining, demonstrating high specificity of staining for sclerostin (Figure S2). Figure 1a shows intense extracellular sclerostin positive granular staining in the stroma of the T. media, forming a linear pattern that tended to align parallel to elastin laminae. Sclerostin staining was discontinuous and occupied approximately 30% of the circumference of the aorta. In contrast to the robust sclerostin staining observed in the T. media of normal human aorta, most (67%) plaques were negative for sclerostin immunoreactivity (presented in detail below) and when present, sclerostin staining in AS plaques was weaker and more variable compared with non-diseased aorta. When sclerostin was detected in plaques, staining was predominantly minimal to mild (semi-quantitative score of 1-2) and typically occupied up to a third of the circumference of the vessel wall, compared to intense (semi-quantitative score of 4) in positive control aorta sections. Sclerostin immunoreactivity was predominantly found in T. media, and sometimes extended into the immediate adjacent subintima (representative image shown in Figure 1b). Sclerostin staining of the subintima also generally occupied up to 30% of the vessel wall circumference and was predominantly minimal to mild in intensity (again, reduced intensity compared to control aorta samples). Importantly, sclerostin staining was never observed in the overlying superficial T. intima, luminal endothelium, associated inflammatory cells (representative image shown in Figure 1c), or within the mineralized/necrotic core of the plaque. Figure 1d shows a diagram summarising the typical location of sclerostin staining in the plaques assessed in this study.

**Figure 1:**
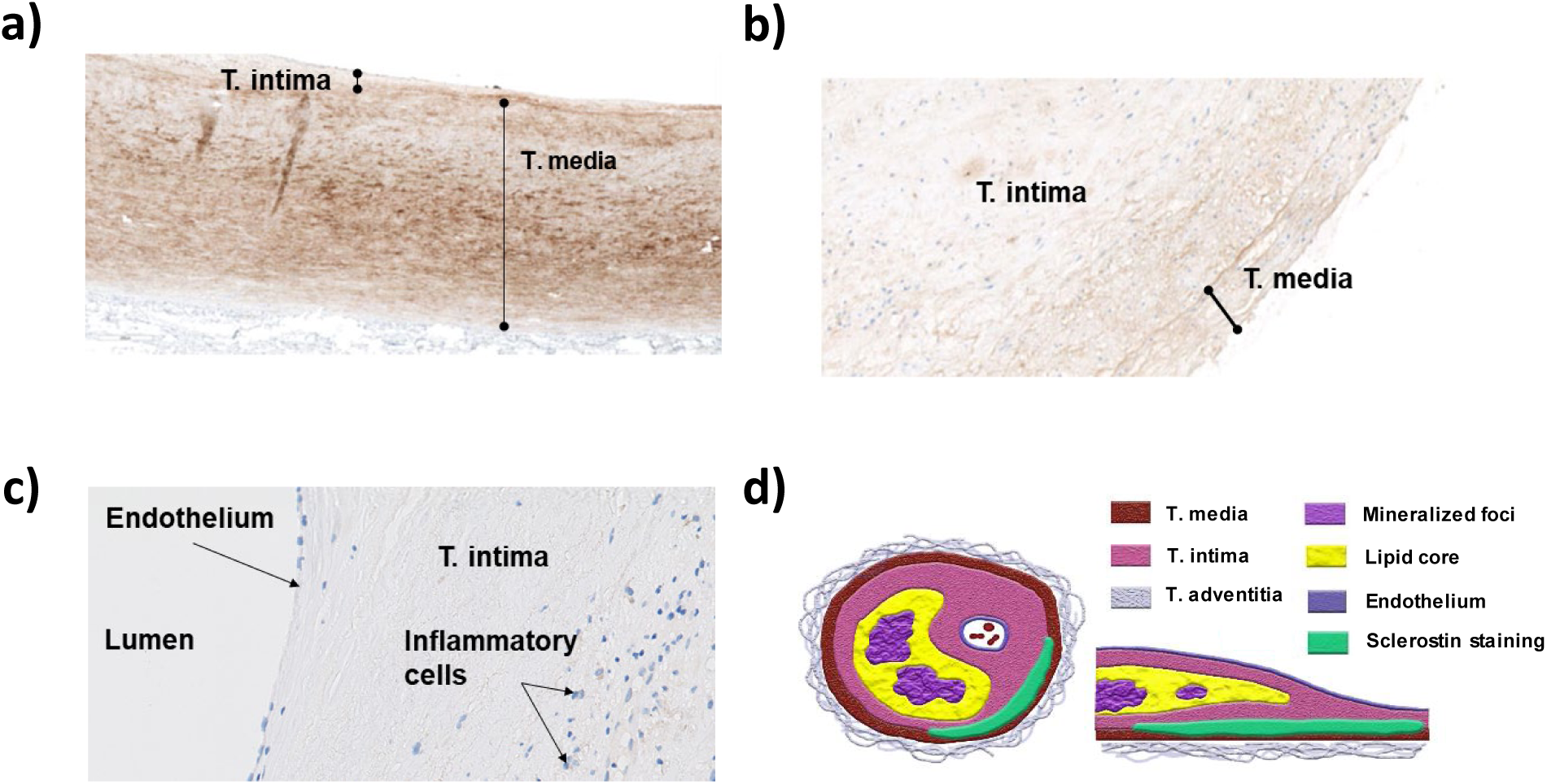
Representative sclerostin staining in human aorta and advanced human AS plaques. a) Intermediate power photomicrograph showing intense sclerostin staining in human aorta; note the strong granular/linear staining within the T media of the aorta orientated parallel to elastin laminae. b) Intermediate power photomicrograph showing mild sclerostin staining in the T. media and immediate adjacent overlying subintima of an advanced human AS plaque. c) Intermediate power photomicrograph showing absence of sclerostin staining in superficial region, endothelium and adjacent inflammatory cells in advanced AS plaque. d) Schematic representation showing histological features of advanced AS plaque and distribution of observed sclerostin staining in transverse and longitudinal section.

Of the plaques surveyed, 67% (97/144) were judged to be negative for sclerostin staining. Fifteen plaques demonstrated staining of both T. media and T. intima (which was always the adjacent subintima), whilst staining of only T. media or only T. intima (which was always deeper regions of subintima distant from the lumen) was recorded in 14 and 18 plaques, respectively. T. media was judged to be absent in 14 of the 144 plaques hence those plaques were not scored for T. media staining. As shown in Figure 2a, the frequency of sclerostin staining was higher in plaques dissected from the carotid artery compared with those removed from the femoral/iliac arteries (42% Vs 16%, respectively). When the location of sclerostin within the plaque was considered, T. media staining was recorded in 29% of carotid plaques Vs 8% of femoral/iliac, whilst the frequencies for T. intima staining were 29% versus 12% in carotid and femoral/iliac plaques, respectively. Due to the relatively low rate of sclerostin staining observed, most subsequent analyses were performed following dichotomisation into “sclerostin staining” or “no sclerostin staining”, based on the simple presence or absence of staining of either T. media or T. intima.

**Figure 2:**
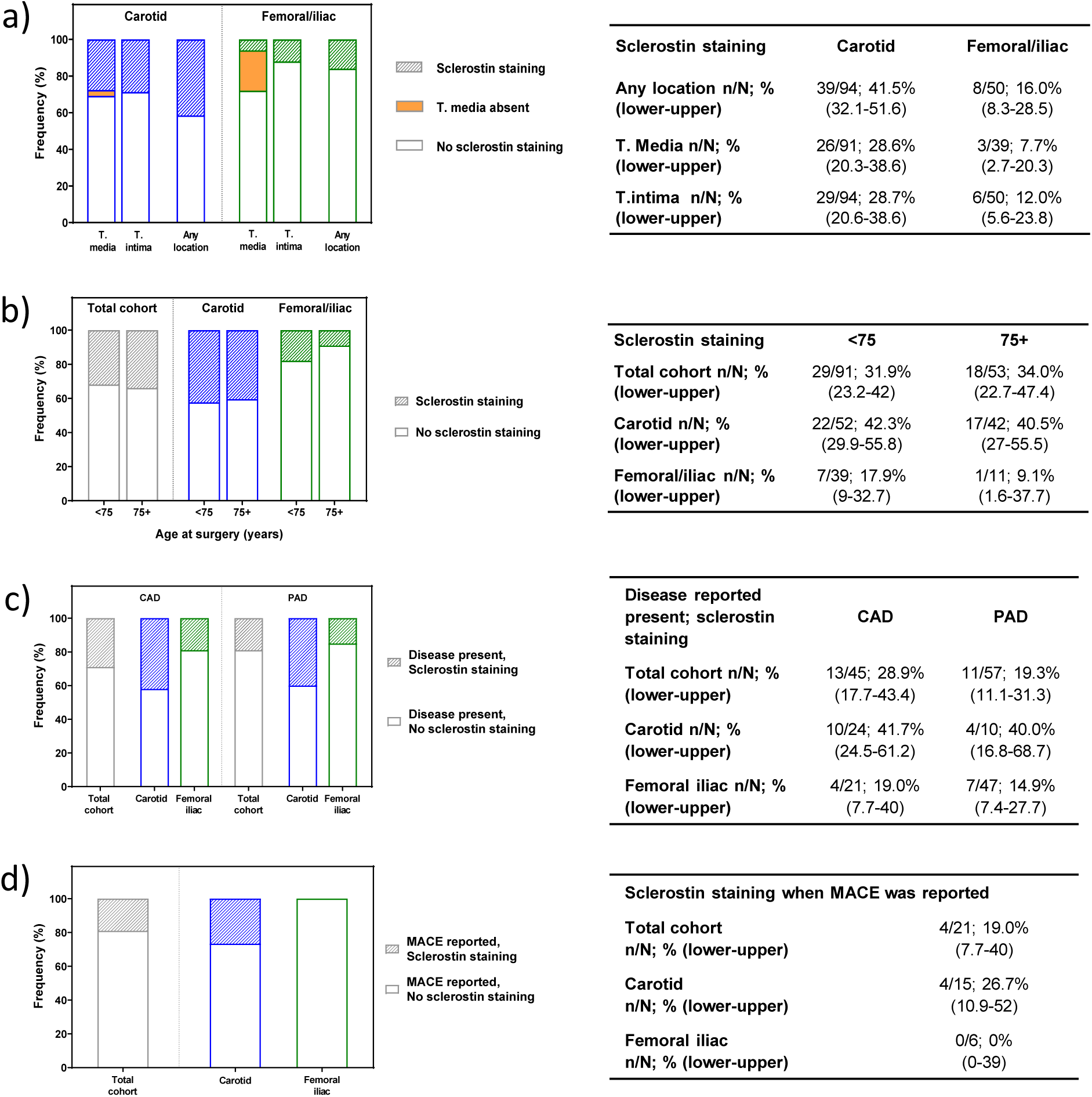
Frequency of sclerostin staining in human AS plaques and association with patient characteristics. a) Frequency of sclerostin staining by plaque anatomical origin (N = 94 carotid; N = 50 femoral/iliac) and location within the plaque. b) Frequency of sclerostin staining by patient age, stratified as less than 75 years (N = 91) or 75 years (N = 53) and older, for plaques collected from any anatomical location or by carotid or femoral/iliac artery subgroups. c) Frequency of sclerostin staining in cohort with a history of CAD (N = 45) or PAD (N = 57) prior to endarterectomy, shown for all plaques and further stratified by anatomical origin of the plaque. d) Frequency of sclerostin staining in subgroup of the total cohort in whom MACE occurred within the 3-year follow-up period (N = 21). Graphs are presented as stacked histograms (shaded bar indicates positive sclerostin staining, empty bar indicates absent sclerostin staining, orange solid bar indicates plaques for which T. media was not present and hence staining of this region could not be scored). Tabulated summaries to the right of each graph presents number of positive sclerostin staining (n) within the cohort (N), percent of positive sclerostin staining within that group with 95% CI shown in brackets.

The risk of major adverse cardiovascular events ((MACE): stroke, myocardial infarction or cardiovascular death) increases with age (30-32). To assess the influence of age on sclerostin staining or endarterectomy location, sclerostin staining was stratified for comparison in women younger than 75 or 75 years and older at the time of endarterectomy. Of the total cohort 63% (91/144) were in the younger group (mean age 63.7 ± 8.2 years, median 67 years) whilst the remaining 37% (53/144) were aged 75 years and above (79.8 ± 3.7 years, median 79 years). The frequency of sclerostin staining in plaques was similar between the age groups (Figure 2b). Approximately half (52/91; 57%) of carotid artery plaques originated from patients less than 75 years of age at the time of surgery and there was no difference in the proportion of carotid artery plaques staining for sclerostin expression by age grouping (42% and 40% for younger and older groups, respectively). In contrast, plaques were more frequently collected from the femoral/iliac arteries of patients in the younger cohort (39/50; 78%) and sclerostin expression was slightly more frequently observed in these samples: 18% compared with 9% staining positive for the older group. Overall, there was no indication of an association between sclerostin staining (presence or absence) and patient age at endarterectomy.

#### 2.1.4. Sclerostin staining association with plaque characteristics and cardiac clinical outcomes

Changes in AS plaque composition have been linked to thrombotic events and thus to clinical outcome (33, 34). The relationship between the sclerostin staining and various features which describe the phenotype and stability of the plaque was assessed (namely intraplaque haemorrhage, lipid content, smooth muscle cells, collagen and calcification; data summarised in Table S2). None of these features showed a statistically significant association with the detection of sclerostin, apart from dystrophic calcification of the acellular necrotic core of plaques, which was inversely correlated with sclerostin staining (p <0.05).

In an ApoE mouse model of atherosclerosis, elevated sclerostin reduced circulating levels of the pro-inflammatory cytokines MCP1, IL6 and TNFα (8), whilst elevation of osteopontin (OPN) in human carotid and femoral plaques was identified as a predictive outcome marker of cardiovascular events (35). This raised the question whether reduced sclerostin would lead to increased pro-inflammatory cytokine levels and/or alter OPN levels. In the current study, there was no linear relationship between sclerostin staining of T. intima or T. media and the expression of MCP1, IL6 and TNFα, or OPN within the subset of plaques for which this data was available (Table S3).

Finally, the clinical history and follow-up data on file was tested for association with patient-reported arterial disease prior to endarterectomy. Coronary artery disease (CAD) was recorded in 31% of the total cohort (45/143; data for one patient was not available), which represented 26% of the carotid artery samples and 42% of the femoral/iliac plaques. Peripheral artery disease (PAD) was noted in 40% (57/143) of the total cohort; the majority (47) of these samples originated from the femoral/iliac group. The presence of arterial disease at surgery did not appear to be associated with sclerostin expression since there was no significant difference in the frequency of positive sclerostin staining in plaques collected from patients with CAD or PAD compared with the total cohort (Figure 2c). MACE (stroke, myocardial infarction or cardiovascular death) occurred in 15% (21/144) of patients during the 3-year follow-up period, with similar rates observed for the carotid and femoral/iliac cohorts (16% in the carotid group Vs 12% in the femoral/iliac group). There was no significant association between sclerostin staining and whether MACE was reported during the 3-year follow-up period (Figure 2d).

### 2.2. Human genetic analyses

To further explore whether a mechanistic lowering of sclerostin levels was associated with increased cardiovascular risk, and could potentially contribute to the imbalance in cardiovascular events observed in the ARCH study, a comprehensive PheWAS was performed. This population level assessment explored whether individuals with lifelong genetically lowered sclerostin expression were at increased risk of cardiac-related outcomes (see Methods). Variants associated with both reduced *SOST* mRNA expression in artery and increased BMD (three variants), or which had been previously associated with reduced circulating sclerostin protein and increased BMD (two variants), were used as a surrogate for non-saturating sclerostin inhibition in participants treated with romosozumab in phase III clinical trials (16, 17, 36). To further explore the validity of the inferences from these variants, four additional variants in the *SOST* region that are associated with BMD were assessed in sensitivity analyses.

#### 2.2.1. Exploration of phenotypes of genetic variants associated with reduced *SOST* expression and sclerostin levels

Three common genetic variants (frequency of the minor allele > 5%) associated with reduced mRNA expression of *SOST* in arteries in GTEx v8 (37) were selected to represent variation across the *SOST* locus; these were rs9899889, rs1107748, rs66838809 (Figure S3 and Table S4); see Methods for detailed selection criteria). These variants cover the three main linkage disequilibrium blocks associated with *SOST* mRNA expression in the region (Figure S4). Alleles associated with lower *SOST* expression in artery (p-values ranging from 7.30 x 10^−11^ to 2.80 x 10^−40^) were positively associated with BMD (overall effect 0.026 (95%CI: 0.023-0.028; p-value = 1.02 x 10^−70^; N = 426,824), Figure S5) and reduced fracture risk (overall effect odds ratio 0.968 (95% CI, 0.959-0.978; p-value = 3.20 x 10^−10^; N cases / N controls = 53,184 / 373,611), Figure S5), thereby indicating a biological outcome consistent with sclerostin pharmacology and positive efficacy data from romosozumab clinical studies.

Whilst these variants were strongly associated with multiple skeletal-related phenotypes (e.g. heel and spine bone mineral density, fracture incidence, standing height), there was no evidence for association of any variants with ischemic or other cardiovascular events following correction for multiple testing (Figure 3a and Table S5; Bonferroni p-value threshold = 0.05 / number of tests performed ≈ 1 × 10^−5^, shown as a dotted line). rs9899889 was associated with lower triglyceride levels and higher HDL cholesterol and apolipoprotein A levels (Tables S6). There is a known genetic association with these lipid measures with the nearby *CD300LG* gene (38-45), in particular with its missense variant rs72836561, and after adjusting the rs9899889 lipid results for rs72836561 there were no longer any significant associations (all p-values > 0.05; Table S6). Moreover, with this adjustment the associations with cardiovascular phenotypes are in-line with what would be expected by chance, with all the circulatory associations either falling within or nearby the 95% confidence interval (94.2% of the associations fall within the confidence interval) in the quantile-quantile plot (Figure 3b(iii)). The circulatory associations are similar to those for all non-skeletal body systems, as shown in Figure 3b(ii), and are in marked contrast to the significant musculoskeletal associations shown in Figure 3b(i), where many associations clearly exceed the dotted line showing the Bonferroni p-value threshold ≈ 1×10^−5^.

**Figure 3:**
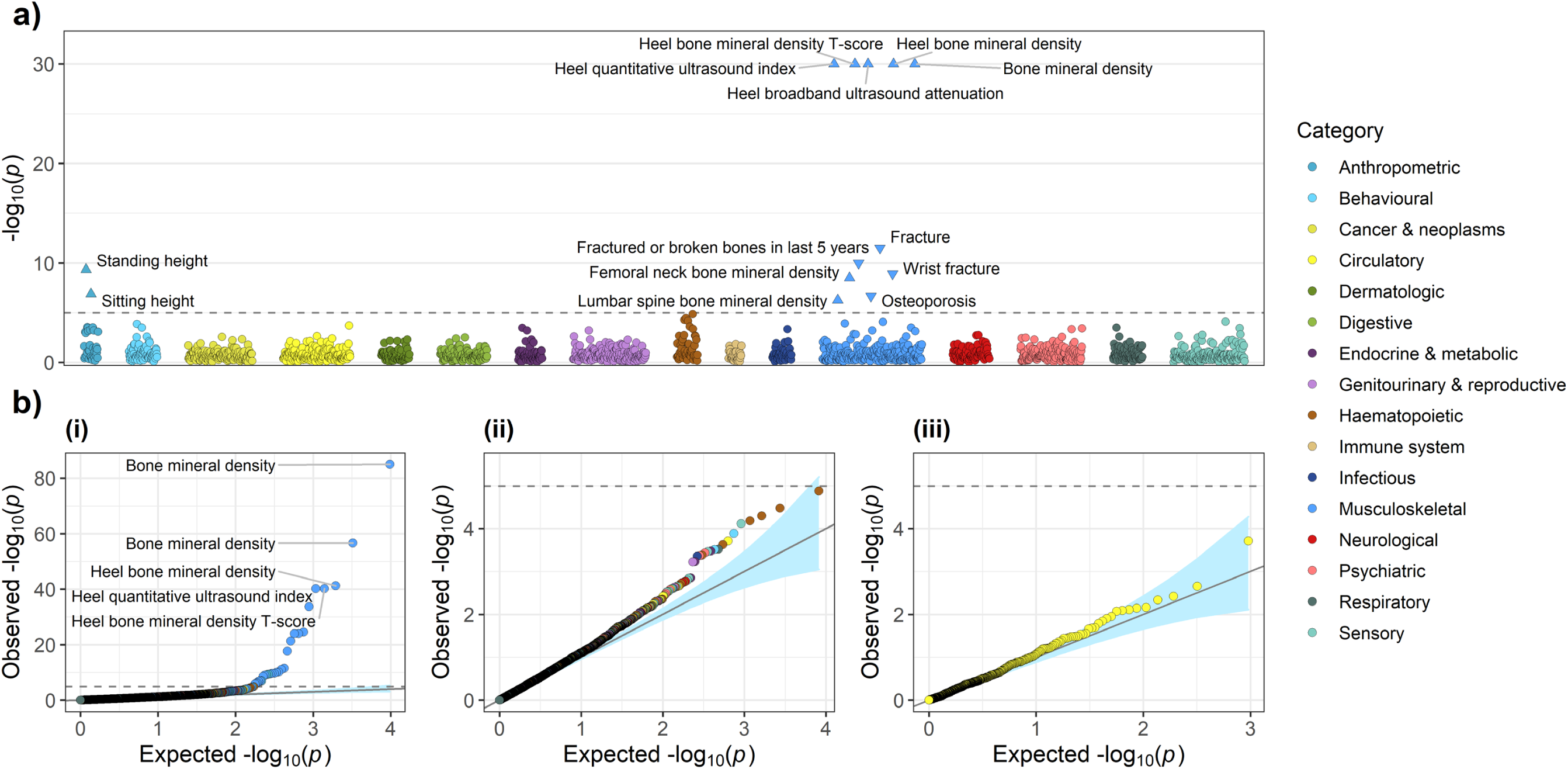
PheWAS of variants associated with reduced *SOST* expression (rs9899889, rs1107748 and rs66838809). a) PheWAS plot of the minimum p-value across the three variants for each phenotype (N phenotypes tested = 1706). b) Quantile-quantile plots for: (i) all associations, (ii) all non-skeletal related associations (i.e. associations not in the anthropometric and musculoskeletal phenotype categories), and (iii) circulatory associations only. Phenotypes were grouped into phenotypic categories. The dashed grey horizontal line indicates a Bonferroni-adjusted p-value threshold of 0.05/*n*_*tests*_ = 0.05/4884 ≈ 1.0 × 10^−5^. Directions of effect for those associations that surpass this threshold are indicated in the PheWAS plot by the direction in which the triangle is pointing. The light blue shaded areas on the quantile-quantile plots represent 95% confidence intervals. The PheWAS plot y-axis was truncated to − log 10 (*p*) = 30. Bone mineral density was estimated using heel ultrasound. Note: the triglyceride, high-density lipoprotein cholesterol and apolipoprotein A results for rs9899889 were replaced with those adjusted for rs72836561 in the *CD300LG* gene (Table S6).

Combining effects across these variants (see Methods 4.2.4 for meta-analysis approach accounting for correlation between variants) for cardiovascular end-points, including coronary artery disease, myocardial infarction and stroke, we found no evidence of association (all p-values > 0.05; Bonferroni p-value threshold of 0.05/18 cardiovascular end-points or risk factors = 0.0028; Figure 4a). Similarly, we found no evidence of association with cardiovascular risk factors, including body mass index, systolic or diastolic blood pressure, low- or high-density lipoprotein cholesterol, or triglycerides (after accounting for the effects of the *CDL300LG* gene), after correcting for multiple testing (Figure 4b). Further inspection of variants in the region surrounding *SOST* for the major cardiovascular endpoints also showed no evidence of genetic associations with these phenotypes (Figure S6).

**Figure 4:**
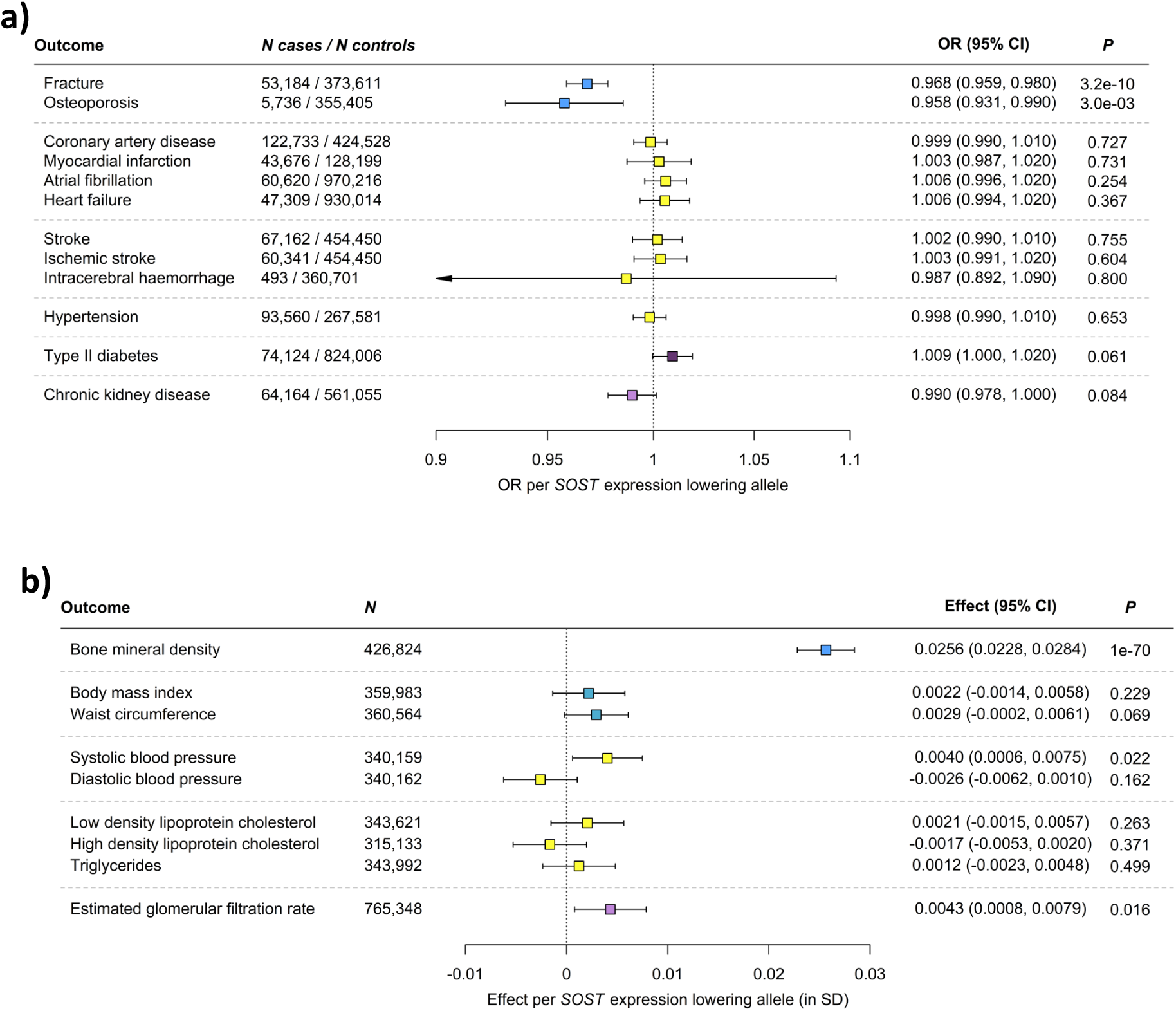
Meta-analysis of variants associated with reduced *SOST* expression and cardiovascular disease end-points and risk factors. a) Associations with disease end-points. b) Associations with bone mineral density and cardiovascular risk factors. Boxes represent point estimates of effects in OR (a) or SD (b) units. Lines represent 95% confidence intervals. The box colouring scheme is the same as that used in Figure 1. Since we tested 18 cardiovascular end-points and risk factors we used a Bonferroni-adjusted p-value threshold of 0.05/*n*_*tests*_ = 0.05/18 = 0.0028. Bone mineral density was estimated using heel ultrasound. Note: the triglyceride and high-density lipoprotein cholesterol results for rs9899889 were replaced with those adjusted for rs72836561 in the *CD300LG* gene (Table S6). The number of samples (N) per outcome is shown within each panel.

We assessed two further variants remote from *SOST* that are associated with reduced circulating sclerostin protein levels in *trans* (46) (rs215226, located on chromosome 12 in *B4GALNT3*; rs7241221, located on chromosome 18 next to *GALNT1*); no *cis* variants were associated with circulating protein levels of sclerostin. These alleles were also associated with increased BMD and other musculoskeletal and anthropometric phenotypes, but were not associated with any cardiovascular phenotypes (Figure S7, Table S7).

#### 2.2.2. Sensitivity analyses using genetic variants associated with BMD in the *SOST* region

Recently, Bovijn *et al*. published a similar set of genetic analyses using variants in the *SOST* region associated with BMD, rs7209826 and rs188810925 (47), and suggested that genetic inhibition of sclerostin could potentially elevate cardiovascular risk (48). Both variants are correlated with the variants selected above: rs7209826 with rs1107748 *r*^2^ = 0.707 and rs188810925 with rs66838809 *r*^2^ = 0.888. For completeness, these variants were also analysed in the current work, and the results were consistent with the variant set already described here; namely, the variants showed positive associations with increased BMD phenotypes, but no associations with any cardiovascular related phenotypes (Figure S8, Table S8). One further additional set of variants in the *SOST* region associated with BMD, rs2741856 and rs7217502 (49) (both variants are correlated with the variants selected above: rs7217502 with rs1107748 *r*^2^ = 0.689 and rs2741856 with rs66838809 *r*^2^ = 0.579), were also analysed and yielded results consistent with all other variants studied (Figure S9, Table S9).

The main inferences made in Bovijn *et al*. are based on a set of fixed-effects meta-analyses, which assume that rs7209826 and rs188810925 are independent. However, these variants are correlated (*r*^2^ = 0.124, *D*’ = 1, p-value = 6.7 × 10^−29^), leading to overly precise results and underestimated p-values. We re-performed their primary analysis of cardiovascular disease events, taking into account the correlation between the two variants (see Methods 4.2.4), and using Bovijn *et al*.’s Bonferroni threshold of 0.004 (main cardiovascular events and risk factors) there is no evidence of association (Figure S10, MI and/or coronary revascularization p-value = 0.011, coronary heart disease (CHD) p-value = 0.078, MACE p-value = 0.019).

## 3. Discussion

This study applied a dual approach to search for potential associations between sclerostin expression in the vasculature and serious cardiovascular events. Atherosclerosis is a driver of most myocardial infarctions and many strokes (18), which are a subset of MACE events. First, immunohistochemistry was used in a cross-sectional study to examine sclerostin expression in FFPE specimens of well-phenotyped AS plaques archived in the AtheroExpress biobank (25). Sclerostin was observed in normal aorta, as expected, but immunoreactivity was low or absent in all plaques examined and most showed no staining. Where present, sclerostin staining was restricted to deeper parts of the plaque/vessel wall (T. media and immediate adjacent subintimal region distant from the lumen) and was of decreased intensity compared to control aorta. In line with these findings, *SOST* down-regulation was reported in human AS plaques compared with aorta (50) and *SOST* expression in the aorta is reduced in rabbit and mouse models of atherosclerosis (15, 51). Sclerostin protein has been detected in human aorta (7), and in the radial artery of patients with advanced chronic kidney disease (13).

Importantly, sclerostin staining was not observed in areas relevant to plaque stability or plaque rupture (i.e. superficial region, fibrous cap, endothelial cells). The nature and characteristics of AS plaques are linked to thrombotic events (23, 34) but sclerostin immunoreactivity was not associated with intraplaque haemorrhage, lipid content, smooth muscle cells or collagen. The inverse association between sclerostin staining and dystrophic calcification of the acellular necrotic core in advanced plaques most likely reflects a passive process of dystrophic mineralisation rather than being an active, cell-mediated or transcriptionally-regulated process (52, 53). Sclerostin staining was also not associated with age at endarterectomy, history of arterial disease prior to, or MACE during the patient follow-up period (minimum 3 years). Finally, sclerostin immunoreactivity was not associated with expression of the pro-inflammatory cytokines MCP1, IL6 and TNFα or with osteopontin, a biomarker of cardiovascular events (35). Based on these findings, it is therefore difficult to postulate a mechanism to explain how inhibition of sclerostin with a sclerostin antibody would have any direct effect on plaque stability.

It is acknowledged that whilst IHC is an ideal technique to examine the location of target protein expression, it is less suited to protein quantitation, hence findings from current study were qualitative. Further, no insights into temporal aspects of sclerostin expression with atheroprogression were gained from this work, although progressive attenuation of *SOST* expression with atheroprogression was reported in a mouse model (15). The total number of sclerostin-positive plaques was also low, which decreased the power to examine associations between sclerostin expression and plaque or patient clinical histories/outcomes.

Immunohistochemistry can yield false positive results. A recent report described IHC staining of sclerostin in AS plaques from patients who underwent carotid endarterectomy (54). In contrast to our findings, Leto *et al*. used a commercially available rabbit polyclonal sclerostin antibody from Abcam and detected sclerostin expression in the entire set of 46 plaques examined, noting immunopositivity in the T. media and core of the plaque, infiltrating macrophages and VSMC. This discrepancy may reflect differences in antibody specificity. In the present study, great care was taken to ensure sclerostin staining was highly specific and sensitive. Comparison of four sclerostin antibodies, including 2 commercially available rabbit polyclonals from Abcam, highlighted the importance of antibody validation, staining optimisation and inclusion of negative control tissues in IHC studies. Unlike the highly specific Scl-Ab #1, which showed no immunoreactivity in tissues known not to express sclerostin protein (i.e. kidney and liver), the commercial reagents exhibited intense staining in both liver and kidney, which suggests non-specific binding (Figure S1) and may account for the apparent widespread “sclerostin” immunoreactivity in atherosclerotic plaques observed by Leto *et al*. (54).

Population level human genetic data can be highly informative in predicting the pharmacological effects of perturbing a drug target, revealing mechanisms of action, identifying alternate indications and predicting adverse drug events (55). To explore whether lowering of sclerostin levels was associated with increased lifetime cardiovascular risk, a comprehensive PheWAS was performed to assess the phenotypic effects of genetic variation simultaneously across thousands of phenotypes from large-scale studies (including UK Biobank, N=361,194). Variants associated with reduced *SOST* expression and increased BMD acted as a proxy for the pharmacological outcome of sclerostin inhibition, albeit reflecting lifelong effects rather than mimicking 12 months treatment with romosozumab.

Here, three different approaches were employed to select *SOST*-related variants for exploration of their association with bone physiology and cardiovascular phenotypes: (i) variants associated with decreased arterial mRNA *SOST* expression and increased BMD, (ii) variants remote from the *SOST* region associated with decreased circulating sclerostin protein (i.e. *trans* acting variants), and (iii) variants in the *SOST* region associated with increased BMD. The results were consistent: *SOST* variants associated with reduced sclerostin expression were associated with increased BMD, reduced risk of fracture and osteoporosis, but were not associated with an increased lifetime risk of any cardiovascular outcomes, including MI, stroke, coronary artery disease, atrial fibrillation, heart failure, or with traits associated with a higher risk such as hypertension and type II diabetes.

The only cardiovascular-related traits showing any association with variants at the *SOST* locus were circulating triglyceride and HDL levels. This association has been reported multiple times previously and attributed to a nearby gene, *CD300LG* (38-45), approximately 100kb upstream of the *SOST* gene. A functional missense variant in *CD300LG* (rs72836561, Arg82Cys) drives the associations in this region (43). After adjusting for the effects of the *CD300LG* gene on triglyceride and HDL levels, there was no evidence of associations with CV traits by more than would be expected by chance. Indeed, when effects across the *SOST* variants were combined, there was no evidence of associations for the major CV endpoints nor with any CV risk factors. *SOST* regional association plots (Figure S6) for the CV endpoints also display the lack of association between variants in this region of the genome with these phenotypes. When the PheWAS analysis was extended to include a range of phenotypic traits across different body systems, there was no evidence that the *SOST* variants were associated with any phenotype other than the expected musculoskeletal and BMD traits (Figures 3, S7, S8 and S9). This consistent lack of association between CV risk and lowered sclerostin using a range of *SOST* variants is compelling since these analyses reflect a lifetime of reduced sclerostin rather than the 12-month duration of sclerostin inhibition in phase III clinical trials with romosozumab. Consistent with the absence of association between reduced *SOST* expression and CV risk factors in this study, there was no evidence of hypertension or alteration in blood pressure, glucose control or lipids, based on treatment-emergent adverse events in clinical trials with romosozumab (56). Furthermore, in nonclinical safety pharmacology studies, there was no evidence of an association between sclerostin inhibition and blood pressure, heart rate or electrocardiograms, and romosozumab or recombinant sclerostin did not elicit vasoconstriction in human coronary rings (15).

A recent paper using similar approaches with two *SOST* variants suggested genetic inhibition of sclerostin could potentially elevate cardiovascular risk (48). It was of interest to understand why the outcome of that analysis differed from the one presented here. Bovijn *et al*. treated their selected variants as if they were independent of each other, however these variants are correlated (p = 6.7 × 10^−29^). This non-independence inflated the precision of associations on all phenotypes studied and underestimated p-values. Indeed, upon accounting for this correlation, and applying a Bonferroni correction for multiple testing, the results for their primary CV disease endpoints were no longer statistically significant (Figure S10). There were also directional discrepancies in the cardiovascular endpoints between biobanks in the supplementary materials of that publication (48), although these could be due to low power in some of these analyses.

It is acknowledged that the genetics approaches in the current study reflect a risk of CV outcomes resulting from a lifetime of genetically lowered sclerostin levels. The comparison between lifelong genetically-driven outcomes with a transient period of sub-saturating sclerostin inhibition is not simple, particularly given the relatively short duration of treatment with romosozumab (12 months), against the cumulative relationship that would be expected with lifelong sclerostin reduction. This is inherent to any study using germline variants as a proxy for pharmacological interventions with significantly shorter observation periods (55). Clinical studies with romosozumab show a rapid non-linear increase in BMD; approximately 65% of the BMD gains in lumbar spine or total hip occurred months 1-6 of the 12-month treatment period (17, 57); in contrast, CV risk gradually increases with age, with an approximately linear increase in risk for each additional decade of life (30). Thus, the genetics approaches may overestimate the impact of reduced sclerostin.

In addition, the variants associated with reduced circulating sclerostin protein were in *trans*, and while *trans-*acting effects of variants affecting expression of genes on other chromosomes have been reported (58), further validation of the relationship between these associations and *SOST* is required. Nonetheless, whilst these variants were associated with increased BMD and other musculoskeletal and anthropometric phenotypes, there was still no evidence of an association between the two trans variants assessed and cardiovascular risk.

The approaches used here utilised common genetic variants. Rare *SOST* variants causing sclerosteosis and van Buchem disease (2, 3, 59) can also inform on the consequence of absent/reduced sclerostin. Sclerosteosis patients typically have a shortened lifespan, largely reflecting complications arising from raised intracranial pressure, but at least 5 have lived for more than 50 years without reported CV effects. Van Buchem patients have a normal lifespan. There are no reports of CV effects in sclerosteosis or van Buchem patients and no mention of CV issues in a published clinical management plan (60, 61).

The data presented here are consistent with the complete nonclinical cardiovascular evaluation of romosozumab, where no functional morphologic or transcriptional effects on the cardiovascular system were observed in animal models in the presence or absence of atherosclerosis (15).

In summary, the data generated from these investigations did not support a mechanistic link between sclerostin inhibition and the imbalance in MI and stroke observed in the phase III ARCH romosozumab study. There was a lack of sclerostin expression in the fibrous cap and luminal endothelium in human arterial atherosclerotic plaques – key sites related to the pathophysiology of plaque rupture and erosion (62) – and a lack of any relationship between sclerostin immunoreactivity and morphological characteristics or biomarkers associated with cardiovascular events. Genetic analyses using common variants associated with reduced expression of the *SOST* gene indicated that natural genetic modulation of *SOST* by variants with a significant positive effect on musculoskeletal readouts, including bone physiology had no significant effect on cardiovascular-related outcomes.

## 4. Methods

### 4.1. Immunohistochemistry of atherosclerotic plaques

#### 4.1.1. Comparison of sclerostin antibody specificity

4μm sections were freshly cut from FFPE blocks, mounted onto SuperFrost Ultra Plus GOLD Adhesion slides (Thermo Scientific) and dried overnight in a 37°C oven. The two proprietary Scl-Abs, Scl-Ab #1 and Scl-Ab #2, were each used at 1:1000 and two commercially available Scl-Abs, ab63097 (Abcam) and ab75914 (Abcam) (referred to as Scl-Ab #3 and #4) were used at 1:900 and 1:50 respectively. The mouse and rabbit isotype matched control antibodies were diluted to give a similar protein concentration to the sclerostin antibodies. Staining was performed using a BOND RX fully automated research stainer (Leica Biosystems) with 20 minutes antigen retrieval (pH6) and 30 minutes incubation with primary antibody. Hematoxylin counterstain was included in all four IHC protocols.

#### 4.1.2. Study design and population

A subset of carotid and femoral/iliac artery plaques from female patients showing 50-95% stenosis who had given informed consent for collaboration with external private partners were selected from the AtheroExpress biobank for the current study.

Positive control tissues included human aorta (one in transverse section, and one in longitudinal section). Human kidney and liver do not express sclerostin protein so were chosen as suitable negative control tissues.

#### 4.1.3. Staining of histological sections

Serial transverse sections (3µm) were freshly cut from FFPE blocks, mounted onto Surgipath X-tra Adhesive micro slides (Leica Biosystems) and maintained at 4°C prior to staining with Scl-Ab #2 (diluted 1:250; approximately 15ug/mL) or isotype matched control antibody (diluted to give a similar protein concentration to sclerostin antibody). Alpha smooth muscle actin antibody (αSMA), Sigma clone 1A4, was used at 1:32000. Staining was performed using Ventana Benchmark Ultra with 12 minutes antigen retrieval (EDTA) and 32 minutes incubation with primary antibody. Hematoxylin counterstain was included in all three IHC protocols.

#### 4.1.4. Visualization and evaluation of stained sections

Following IHC staining, slides were visualized under light microscopy by the study pathologist for expression of sclerostin protein. The study pathologist judged each slide for adequacy and quality of tissue elements, and specificity of staining before assessing sclerostin expression within each plaque. The intensity, percentage circumference of staining and region of staining was assigned as applicable.

Interpretation was based on comparisons to physiologic/expected sclerostin expression and staining patterns/histologic locations within positive control aorta samples. The presence of granular deposits arranged in a linear or semi-linear pattern, parallel to stromal elements was considered positive sclerostin staining, consistent with a secreted protein from vascular smooth muscle cells (VSMCs). Non-specific staining was associated with section lifting/folding and often produced characteristic geometric patterns distributed randomly within the sample. Control article, αSMA and H&E slides for each tissue were reviewed by the study pathologist. These were used for adjunctive comparison with the sclerostin IHC-stained slides and to identify histological features within the plaques.

#### 4.1.5. Sclerostin IHC scoring

Two positive control aorta samples were scored for staining intensity. One transverse aorta section was scored for circumferential percentage staining whereas only location and staining characteristics were described for the other longitudinal section. All plaque samples were scored for sclerostin staining intensity, circumferential percentage staining, and for the T. intima only, location of staining (superficial, mid or deep) using the following criteria:

- Intensity of staining was scored semi-quantitatively on a scale of 0-4 where: 0=no staining; 1=minimal pale staining that could clearly be distinguished from non-specific background staining; 2=mild staining that was more intense than minimal; 3=moderate staining that was more intense than mild; 4=severe staining that was similar in intensity noted in positive control aorta samples and of dark brown-black coloration
- Circumferential staining was scored semi-quantitatively to the nearest whole unit of 10 as a percentage of total circumference identified under low power magnification. Where staining was <10% this was indicated as an approximate % score and/or described in the comments
- In the T. intima, staining location was scored on the following 5 levels: Level 1=superficial plaque/T. intima in area of plaque cap; Level 2=mid region of plaque between Level 1 and 3; Level 3=deep plaque/T. intima adjacent to T. media; Level 4=mid and deep region of plaque/T. intima (i.e. Level 2 & Level 3); Level 5=superficial, mid and deep region of plaque/T. intima

#### 4.1.6. Integration of sclerostin staining with AtheroExpress biobank data

Due to the relatively small number of samples showing sclerostin staining, the cohort was dichotomized into two groups; one group without sclerostin staining, and one group with sclerostin staining. Analyses included the proportion of plaques in which sclerostin expression was observed in relation to baseline patient characteristics, namely age at endarterectomy and recorded history of coronary artery disease or peripheral artery disease. The association of sclerostin staining with the major adverse cardiovascular event (MACE: stroke, myocardial infarction or cardiovascular death) endpoint was also examined.

#### 4.2. Human genetic analyses

#### 4.2.1. *SOST* genetic variant selection

To select genetic variants that mimic *SOST* inhibition variants associated with lower mRNA *SOST* expression were chosen. All variants that satisfied the following criteria were eligible for selection:

- Associated with mRNA *SOST* expression in tibial artery or heart tissue in GTEx v8 (37, 63) (as cardiovascular endpoints are of interest here and as bone is not included in GTEx) at genome-wide significance level (5×10^−8^) (63)
- Associated with BMD levels, estimated using heel ultrasound, at genome-wide significance level (49)
- Within ±100kb of the *SOST* gene

These eligible variants (of which there were 99) were then linkage disequilibrium (LD) pruned using LDlink (*r*^2^ cut-off of 0.2) (64) using the European samples from 1000 Genomes, a whole-genome sequencing project commonly used in the genetics field to compute LD statistics, as the LD reference panel (65). The same variants were selected if the analysis was restricted to tibial artery only and if results from all tissues in GTEx v8 were used.

Two additional strategies were used to select variants related to reduced sclerostin levels. The first are variants that are associated with circulating sclerostin protein levels at genome-wide significance level (rs215226 & rs7241221) (46) and the second are two variant sets associated with BMD levels in the *SOST* region (rs7209826 & rs188810925 and rs2741856 & rs7217502) found in recent large-scale genome-wide association studies (47, 49). The correlations between the variants in these variant sets are displayed in Figure S4.

Correlation statistics between *SOST* variants, i.e. LD statistics, were computed using the European samples from 1000 Genomes (65). The measures computed were (similar to Pearson’s correlation coefficient) and ′ (66). A chi-squared test of independence (for a 2 × 2 table) was used to assess the relationship between two variants.

#### 4.2.2. Phenome-wide association study

The PheWAS was performed using look-ups from the latest UK Biobank genetic analyses (mean age = 56.5 years and proportion of males = 46.2%) from the Neale Lab (www.nealelab.is/uk-biobank/ and (67)) and PhenoScanner (68, 69), as well as from additional datasets related to bone physiology and cardiovascular disease and risk factors (49, 70-76). For completeness, the variants were also queried in the GWAS Catalog (77) and IEU GWAS server (78), but no additional relevant associations were identified. The full list of studies used in the present analysis are available in Table S10. Of particular interest are those datasets related to bone physiology and cardiovascular disease. The genetic associations with these phenotypes were studied in large population-based studies: GEFOS (bone mineral density N = 426,824, fracture risk N cases / N controls = 53,184 / 373,611 (49)), CARDIoGRAMplusC4D (coronary artery disease N cases / N controls = 122,733 / 424,528 (75), myocardial infarction N cases / N controls = 43,676 / 128,199 (73)) and MEGASTROKE (all stroke N cases / N controls = 67,162 / 454,450, ischemic stroke N cases / N controls = 60,341 / 454,450 (71)).

To ensure that one association is included in the analysis for each variant-phenotype pair, we selected a single result for each pair based on the largest number of cases for binary phenotypes and the largest number of samples for continuous phenotypes. All phenotypes were categorised into one of the following phenotypic categories: Anthropometric, Behavioural, Cancer & neoplasms, Circulatory, Dermatologic, Digestive, Endocrine & metabolic, Genitourinary & reproductive, Haematopoietic, Immune system, Infectious, Musculoskeletal, Neurological, Psychiatric, Respiratory and Sensory.

#### 4.2.3. Conditional analyses

Since there is a well-known genetic association between *CD300LG* and triglyceride and HDL levels in the *SOST* gene region, we used conditional analyses to adjust for genetic variations related *CD300LG*. As apolipoprotein A is highly correlated with HDL levels (genetic correlation = 0.91), we also adjusted these associations for variations related *CD300LG*. These conditional analyses were performed using the COJO methodology (81), which allows variant effects to be adjusted for each other using summary level results. The European samples from 1000 Genomes were used as the LD reference panel (65). If a nominal association of p<0.001 (at the PheWAS level) was observed for at least one of these lipid measures with a *SOST* variant, these associations for this variant were either adjusted for rs72836561 (missense variant in *CD300LG*) or rs72836567 (downstream of *CD300LG* and the top associated variant with mRNA *CD300LG* expression (37)), where the choice of *CD300LG* variant was based on the correlation structure with the *SOST* variant.

#### 4.2.4. Meta-analysis of allelic effects

Effects across variants were meta-analysed for key clinical endpoints and risk factors using fixed-effects meta-analysis accounting for any residual correlation between the variants (82, 83). This model is a weighted generalized linear regression of the form 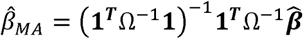 and 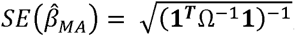, where 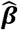 is the vector of variant-phenotype estimates, **1** is an intercept vector and Ω= ***σσ*** ^*T*^ ° ***ρ***, where *σ* is the vector of corresponding standard errors of 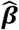 and ***ρ*** is the variant correlation matrix. The genotype correlation statistics between the variants (that make up ***ρ***) were computed using the European samples from 1000 Genomes (65).

### 4.3. Statistics

For the immunohistochemistry study: both histological plaque features and subsequent cardiovascular events were tested for an association with sclerostin staining using Pearson Chi-Square test. A linear regression model was used to test for a linear relationship between sclerostin staining and the concentration of cytokines (IL6, MCP1, TNFα) and osteopontin (OPN) measured in plaque. These cytokines were selected based on the availability of pre-existing data in the AtheroExpress biobank. These statistical analyses were performed using SPSS version 21.0 (SPSS Inc., Chicago, IL) and a p-value of <0.05 was considered significant.

For the human genetic analyses: In PheWAS, many variant-phenotype associations were tested, therefore a Bonferroni adjusted p-value of 0.05 divided by the number of tests performed was used to account for multiple testing (e.g. in the primary analysis of variants associated with *SOST* mRNA expression the threshold = 0.05/4884≈1.0×10^−5^) (79). All associations with binary phenotypes on the risk difference scale were transformed to the odds ratio scale (80). In the analysis of cardiovascular endpoints and risk factors,18 phenotypes were assessed, therefore a Bonferroni-adjusted p-value threshold of 0.05/18=0.0028 was used. The genetic data processing and analyses were performed using python version 3.6 (package: pandas (84)), R version 3.6 (packages: ggplot2 (85), pheatmap and metafor (86)) and tabix (87).

### 4.4. Study approval

Carotid and femoral/iliac artery plaques from the AtheroExpress biobank were collected from patients who had given written informed consent as previously described (25); only plaques from patients who had given informed consent for collaboration with external private partners were selected for the current study.

All the participants in the UK Biobank, GEFOS, CARDIoGRAMplusC4D and MEGASTROKE cohorts provided written informed consent for participating in research studies. Blood or saliva samples were collected according to protocols approved by local institutional review boards. Details are provided in the original publications describing the cohorts (49, 67, 71, 73, 75).

## Data Availability

Datasets generated during and/or analysed during this study which were not included as supplementary files are available from the corresponding author on reasonable request.

## Supplemental tables in Excel file

Table S1: Baseline characteristics of AtheroExpress patients included in this study.

Table S2: Association of sclerostin staining with plaque features.

Table S3: Association of sclerostin staining with cytokine expression in plaques

Table S4: The *SOST* variants selected using mRNA *SOST* expression, sclerostin protein levels and BMD levels in the *SOST* region and their associations with mRNA *SOST* expression, BMD levels and fracture risk.

Table S5: PheWAS results for variants associated with reduced *SOST* expression and increased BMD in the *SOST* region (rs9899889, rs1107748 and rs66838809).

Table S6: *SOST* variant associations with triglyceride, HDL and Apolipoprotein A levels adjusted for variants related to the *CD300LG* gene.

Table S7: PheWAS results for variants associated with reduced sclerostin protein levels (rs215226 and rs7241221).

Table S8: PheWAS results for variants associated with increased BMD in the *SOST* region (rs7209826 and rs188810925).

Table S9: PheWAS results for variants associated with increased BMD in the *SOST* region (rs2741856 and rs7217502).

Table S10: List of genetic association studies used in the PheWAS analysis.

## Author contributions

GH, PH, RO, RWB, JRT and GP conceptualised and designed the IHC study; RO performed the Scl-Ab specificity and optimisation experiments; PH, IVK, GH and GP analysed the IHC study data. GH, JRS, CV, AW and MA conceptualised the genetics experiments; JS and CV performed human genetic association studies and analysed the data. GH, JS, PH, CV and AW wrote the manuscript; all co-authors reviewed and approved the final version of the submitted manuscript.

## Acknowledgements

The authors would like to thank Dr Chris Paszty (Amgen Inc.), Dr Jochen Dunkel (UCB Pharma) for their helpful support, and Petra van der Kraak (University Medical Centre Utrecht) for technical support to perform IHC staining of AS plaques.

Summary statistics were downloaded from the NHGRI-EBI GWAS Catalog (77) for studies GCST006979, GCST006980 (49), GCST005194 (75), GCST005838, GCST005843 (71), GCST006414 (72), GCST009541 (74), GCST007517 (70), GCST008058, GCST008064 (76), downloaded on 25/06/20. Data on coronary artery disease and myocardial infarction have been contributed by CARDIoGRAMplusC4D investigators and have been downloaded from www.cardiogramplusc4d.org. The MEGASTROKE project received funding from sources specified at http://www.megastroke.org/acknowledgments.html. We thank the MEGASTROKE authors, the author list is available here: https://www.megastroke.org/authors.html. All summary genetic association results used are available in the public domain.

