## Supplementary Figures for "Genetic and atherosclerotic plaque immunohistochemical analyses do not associate reduced sclerostin expression with cardiovascular events"

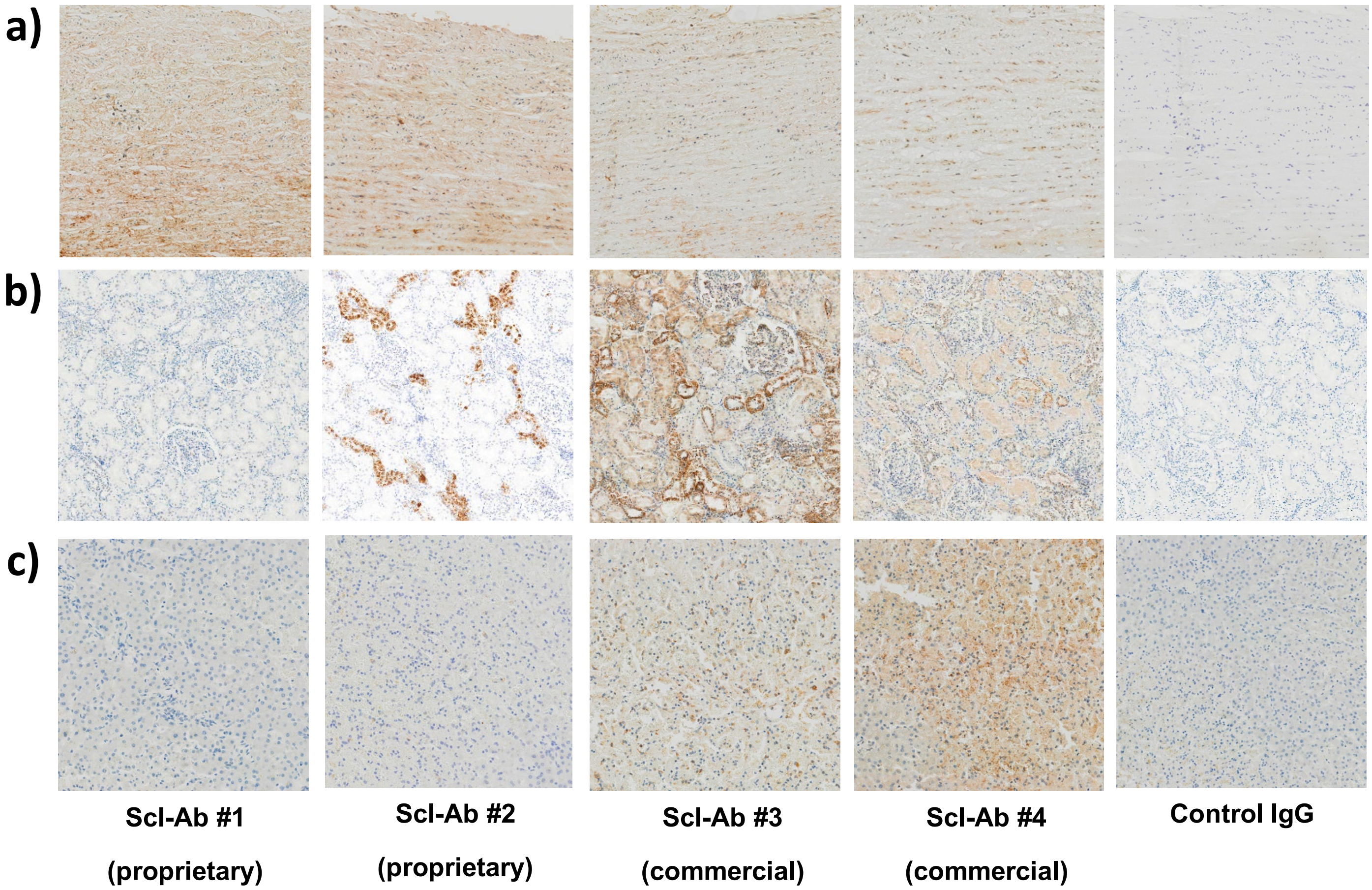


**Figure S1: Evaluation of sclerostin antibody specificity in immunohistochemical staining of FFPE tissue sections.**

Two proprietary and two commercially-available sclerostin antibodies were tested for their ability to sensitively and specifically stain sclerostin protein in FFPE tissue sections (a, aorta; b, kidney; c, liver). All 4 antibodies tested showed strong staining of aorta T. media and 3 antibodies exhibited background staining in other tissues where sclerostin is not expressed. Only one of the antibodies tested (Scl-Ab #1) showed both specific staining of positive control tissues, and absent staining of the negative control section; this antibody was used to stain the 144 AS plaques selected from the AtheroExpress Biobank.

Images are shown at 10x magnification.

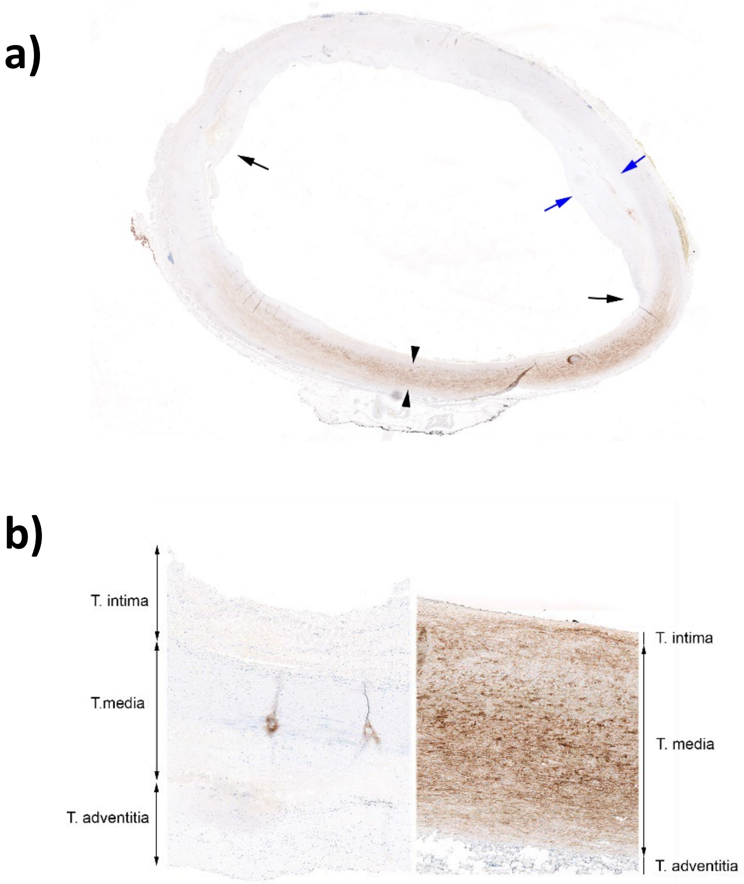


**Figure S2: Representative sclerostin staining of human aorta.**

a) Low power sub-gross photomicrograph of human aorta showing regional distribution of sclerostin immunohistochemical staining. Blue arrows indicate intimal thickening (early atherosclerotic plaque), black arrows indicate shoulder region and black arrowheads positive staining in T. media. Note that sclerostin staining is downregulated at shoulder region and absent beneath early plaque. b) Low power photomicrograph of human aorta showing an IgG isotype-control stained negative control (left) and sclerostin stained positive control (right). No specific staining is present in the control IgG-stained section. The sclerostin stained section showed strong (severe, grade 4) staining in T. media.

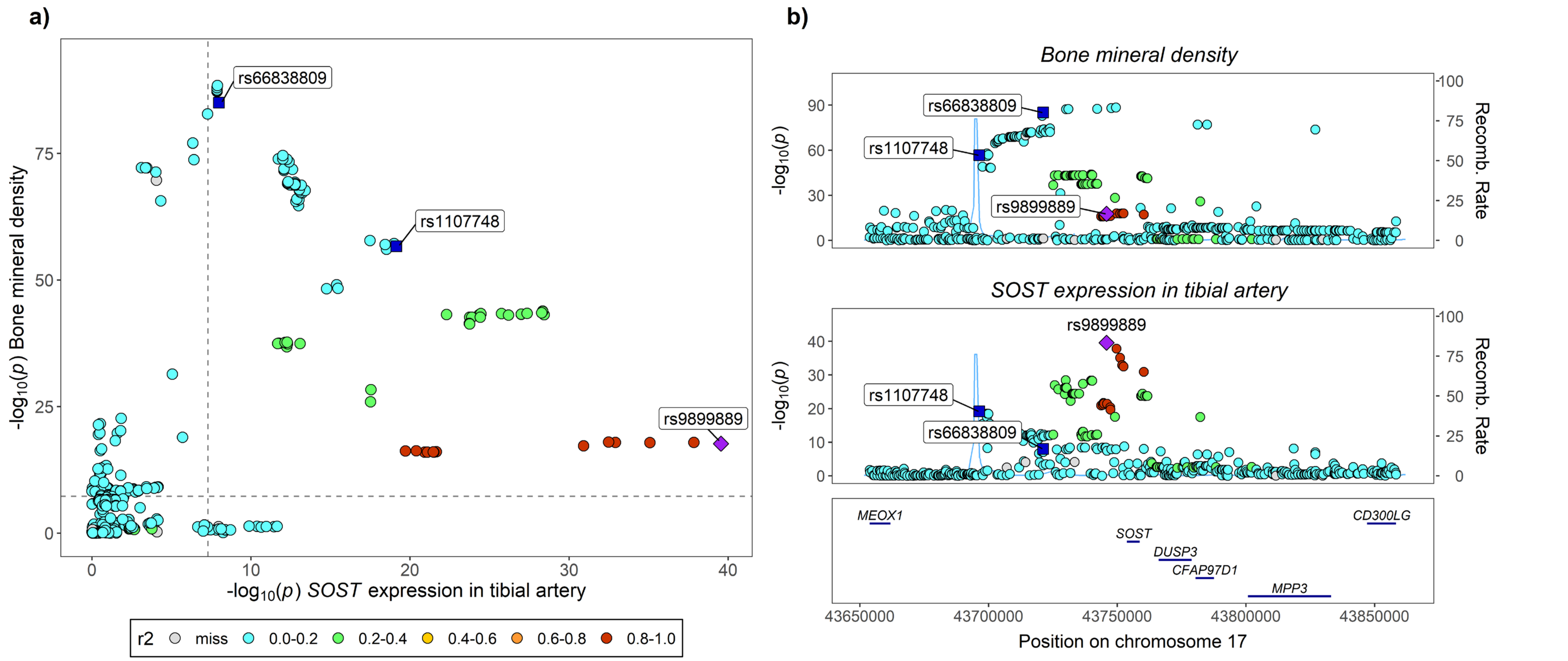


**Figure S3: Locus comparison between *SOST* mRNA expression in tibial artery and BMD levels.** a) Plot of the p-values for *SOST* mRNA expression against the p-values for BMD levels. b) stacked regional association plots of *SOST* mRNA expression in tibial artery and BMD levels using gassocplot2. Each point represents a p-value, transformed using the -log10, for a variant and the colours of the points indicate the linkage disequilibrium (in $r^2$) of each SNP with rs9899889 (indicated as a purple point; rs1107748 and rs66838809 are also highlighted as blue squares). The genomic window plotted is ±100kb either side of the *SOST* gene based on build 38 coordinates. The horizontal and vertical dashed grey lines in the locus compare plot indicate the genome-wide significance threshold ($5 \times 10^{-8}$). Bone mineral density was estimated using heel ultrasound.

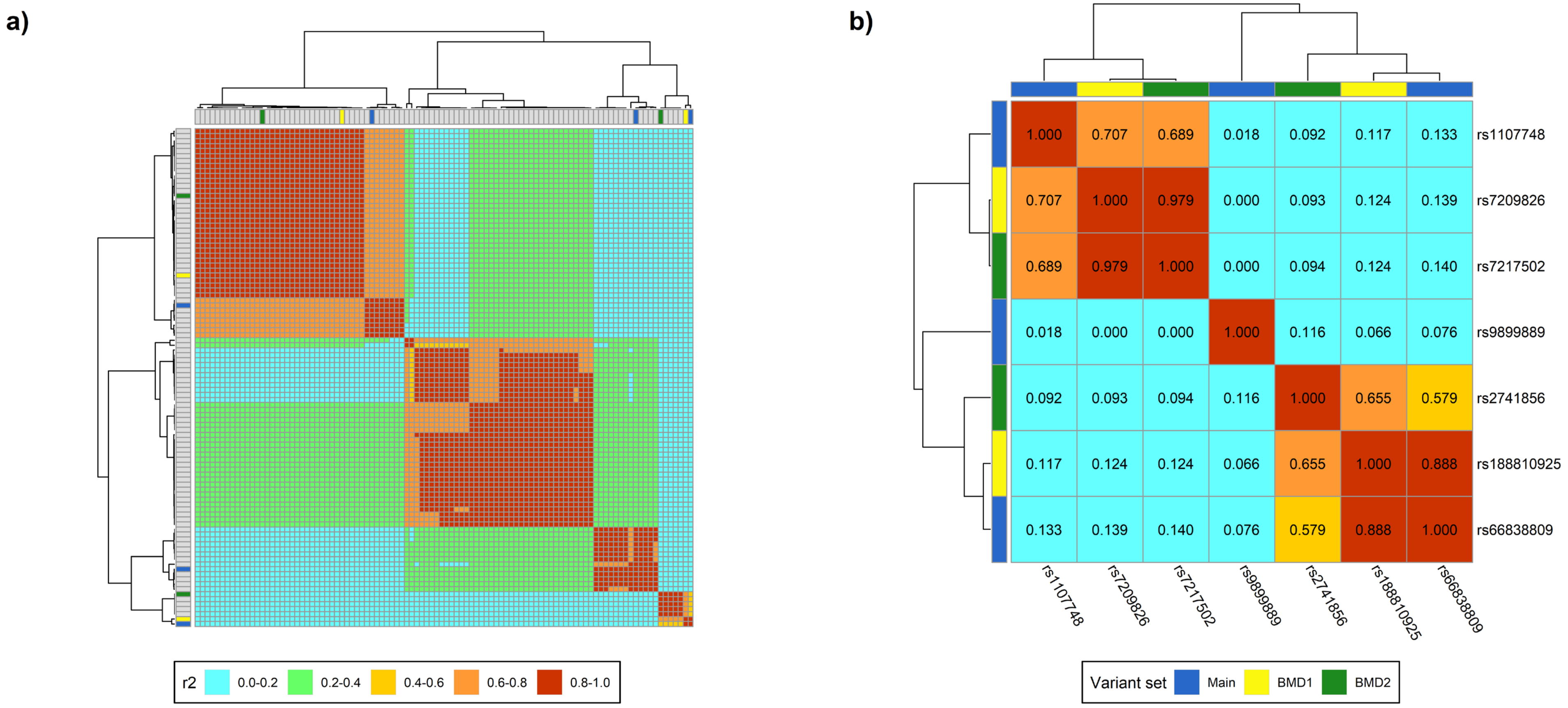


**Figure S4: Linkage disequilibrium between the *SOST* variants.**

a) Variants that have an association with both *SOST* mRNA expression and bone mineral density in the *SOST* region (±100kb either side of the *SOST* gene).

b) The variant sets from the *SOST* region used in the analysis. The variant sets are those used in the primary analysis (Main: rs9899889, rs1107748 & rs66838809) and the two BMD variant sets (BMD1: rs7209826 & rs188810925; BMD2: rs2741856 & rs7217502).

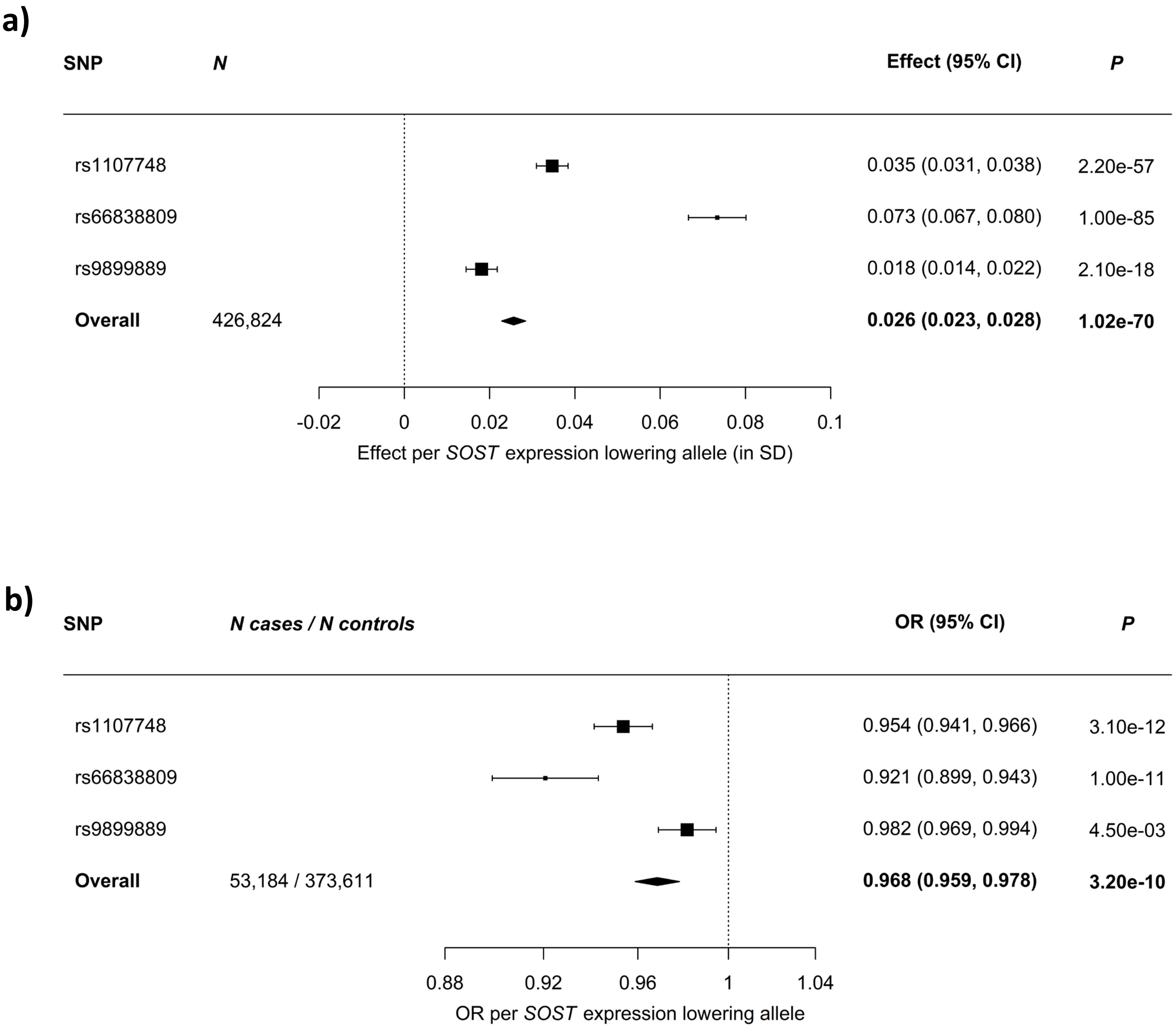


**Figure S5: Meta-analysis of variants associated with reduced *SOST* expression with bone mineral density and fracture risk.**

a) Associations with bone mineral density. b) Associations with fracture risk. Boxes represent point estimates of effects in SD (a) or OR (b) units. Lines represent 95% confidence intervals. Bone mineral density was estimated using heel ultrasound.

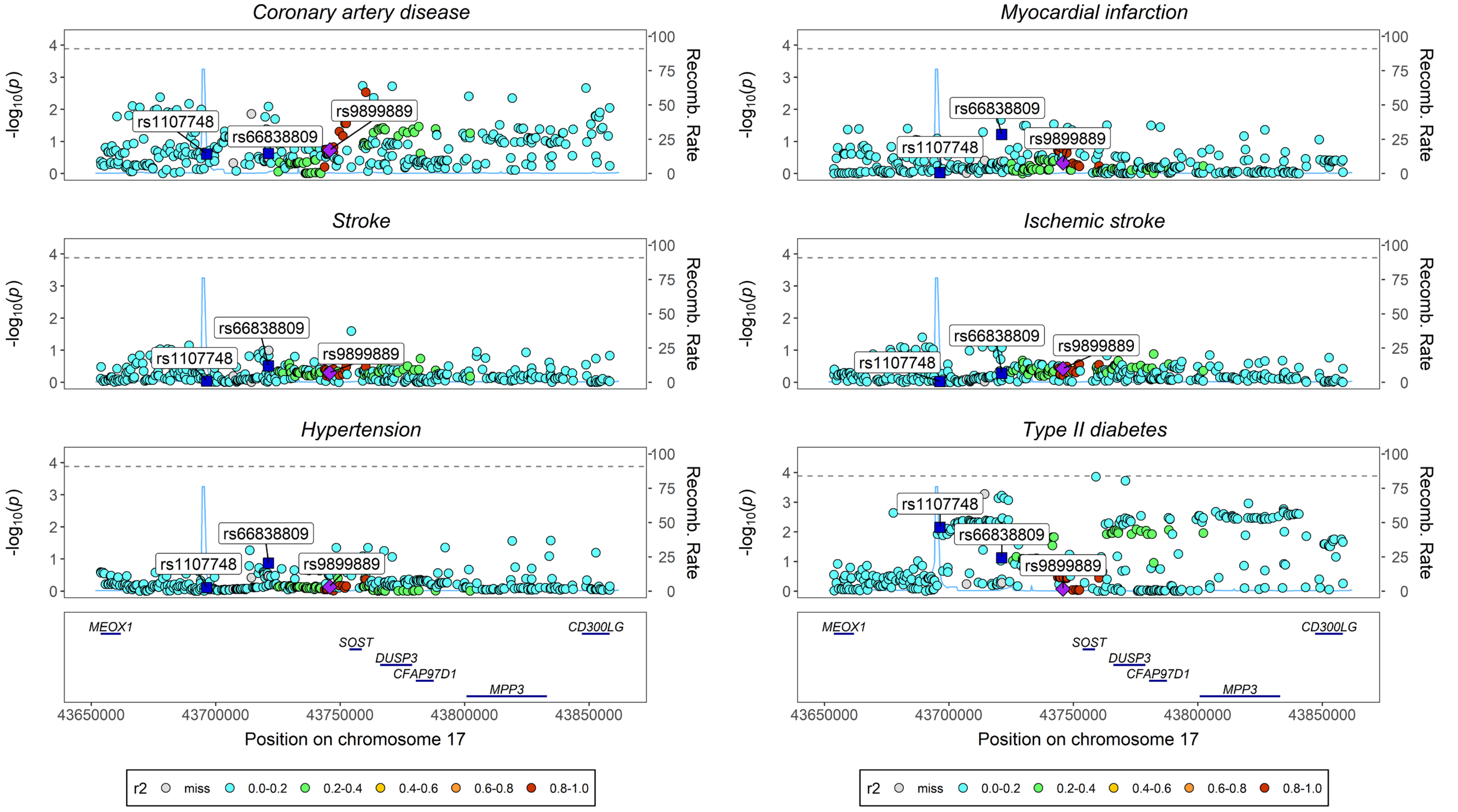


**Figure S6: Regional association plots of the *SOST* region for cardiometabolic end-points.**

The plots are for coronary artery disease, myocardial infarction, all stroke, ischemic stroke, hypertension and type II diabetes. Each point represents a p-value, transformed using the -log10, for a variant and the colours of the points indicate the linkage disequilibrium (in $r^2$) of each SNP with rs9899889 (indicated as a purple point; rs1107748 and rs66838809 are also highlighted as blue squares). The genomic window plotted is ±100kb either side of the *SOST* gene based on build 38 coordinates. The dashed grey horizontal line indicates a Bonferroni-adjusted p-value threshold of $0.05/n_{variants} = 0.05/382 \approx 1.3 \times 10^{-4}$. This plot was created using gassocplot2.

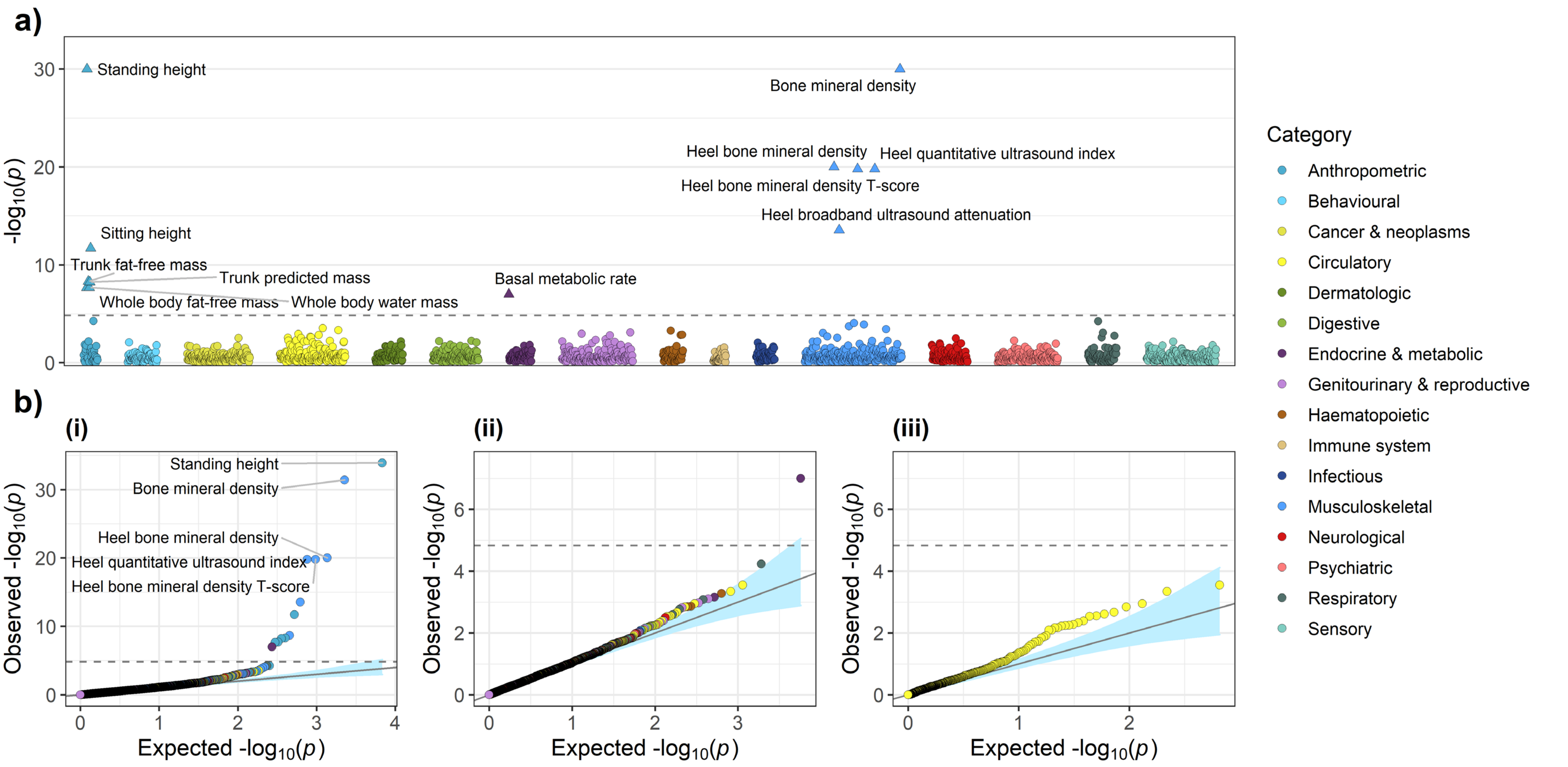


**Figure S7: PheWAS of variants associated with reduced sclerostin protein levels (rs215226 and rs7241221).**

a) PheWAS plot of the minimum $p$-value across the three variants for each phenotype. b) Quantile-quantile plots for: (i) all associations, (ii) all non-skeletal related associations (i.e. associations not in the anthropometric and musculoskeletal phenotype categories), and (iii) circulatory associations only. Phenotypes were grouped into phenotypic categories. The dashed grey horizontal line indicates a Bonferroni-adjusted p-value threshold of $0.05/n_{tests} = 0.05/3408 \approx 1.5 \times 10^{-5}$. Directions of effect for those associations that surpass this threshold are indicated in the PheWAS plot by the direction in which the triangle is pointing. The light blue shaded areas on the quantile-quantile plots represent 95% confidence intervals. The PheWAS plot y-axis was truncated to $-\log 10\,(p) = 30$. Bone mineral density was estimated using heel ultrasound.

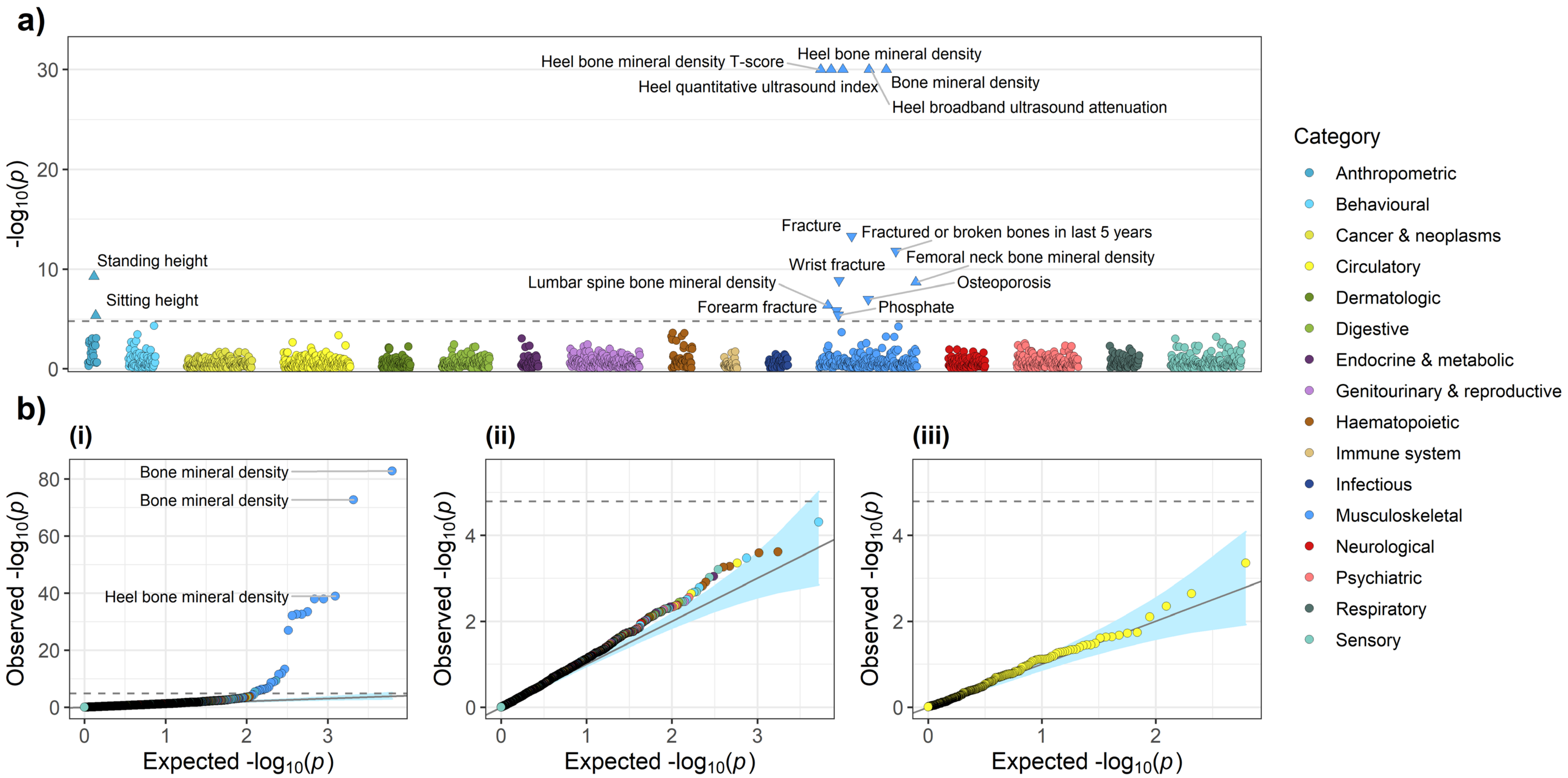


**Figure S8: PheWAS of variants associated with increased BMD in the *SOST* region (rs7209826 and rs188810925).**

a) PheWAS plot of the minimum $p$-value across the three variants for each phenotype. b) Quantile-quantile plots for: (i) all associations, (ii) all non-skeletal related associations (i.e. associations not in the anthropometric and musculoskeletal phenotype categories), and (iii) circulatory associations only. Phenotypes were grouped into phenotypic categories. The dashed grey horizontal line indicates a Bonferroni-adjusted p-value threshold of $0.05/n_{tests} = 0.05/3100 \approx 1.6 \times 10^{-5}$. Directions of effect for those associations that surpass this threshold are indicated in the PheWAS plot by the direction in which the triangle is pointing. The light blue shaded areas on the quantile-quantile plots represent 95% confidence intervals. The PheWAS plot y-axis was truncated to $-\log 10\,(p) = 30$. Bone mineral density was estimated using heel ultrasound. Note: the triglyceride, high-density lipoprotein cholesterol and apolipoprotein A results for rs7209826 were replaced with those adjusted for rs72836567 near the *CD300LG* gene (Table S6).

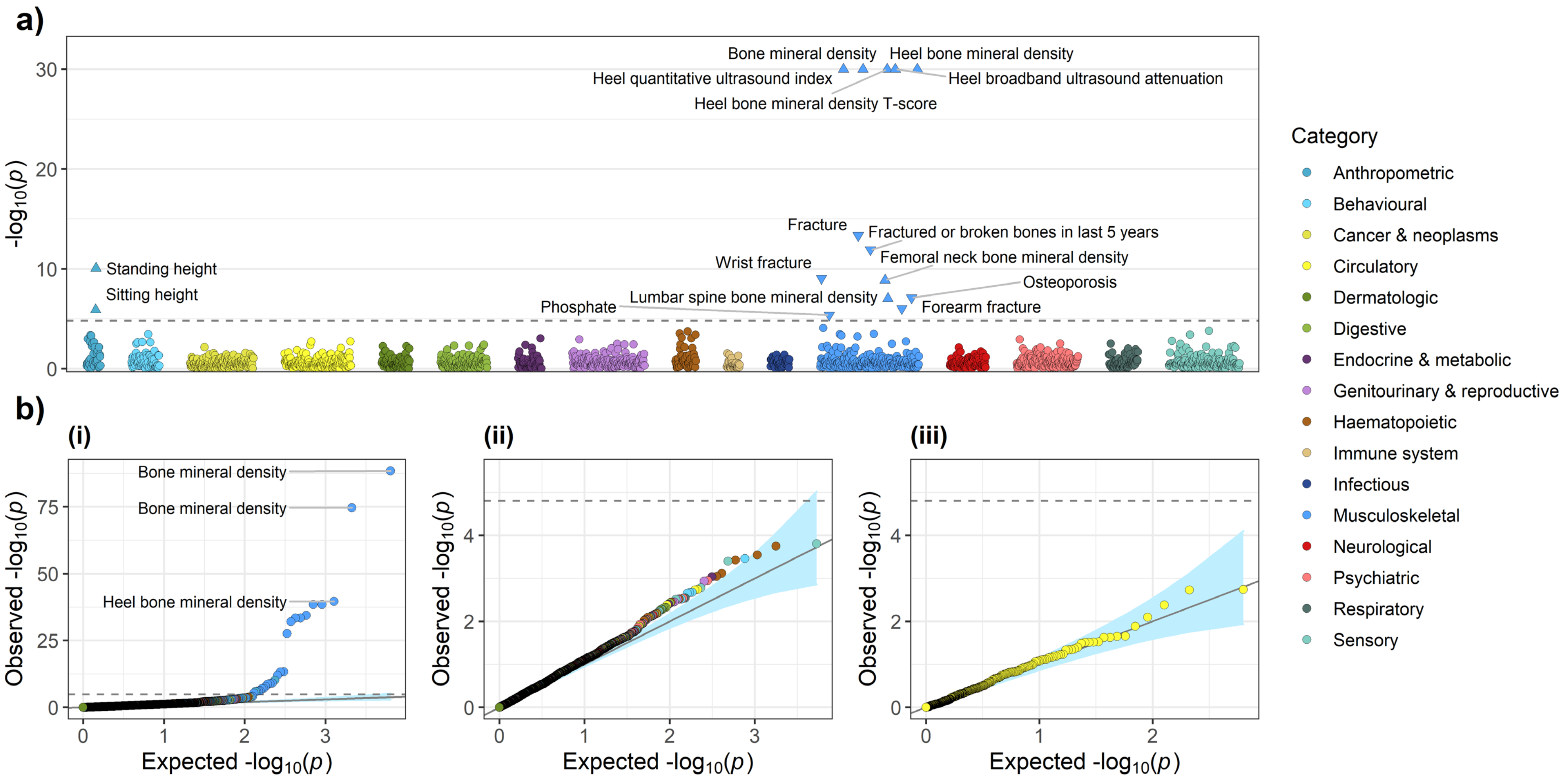


**Figure S9: PheWAS of variants associated with increased BMD in the *SOST* region (rs2741856 and rs7217502).**

a) PheWAS plot of the minimum $p$-value across the three variants for each phenotype. b) Quantile-quantile plots for: (i) all associations, (ii) all non-skeletal related associations (i.e. associations not in the anthropometric and musculoskeletal phenotype categories), and (iii) circulatory associations only. Phenotypes were grouped into phenotypic categories. The dashed grey horizontal line indicates a Bonferroni-adjusted p-value threshold of $0.05/n_{tests} = 0.05/3184 \approx 1.6 \times 10^{-5}$. Directions of effect for those associations that surpass this threshold are indicated in the PheWAS plot by the direction in which the triangle is pointing. The light blue shaded areas on the quantile-quantile plots represent 95% confidence intervals. The PheWAS plot y-axis was truncated to $-\log 10\,(p) = 30$. Bone mineral density was estimated using heel ultrasound. Note: the triglyceride, high-density lipoprotein cholesterol and apolipoprotein A results for rs7217502 were replaced with those adjusted for rs72836567 near the *CD300LG* gene (Table S6).

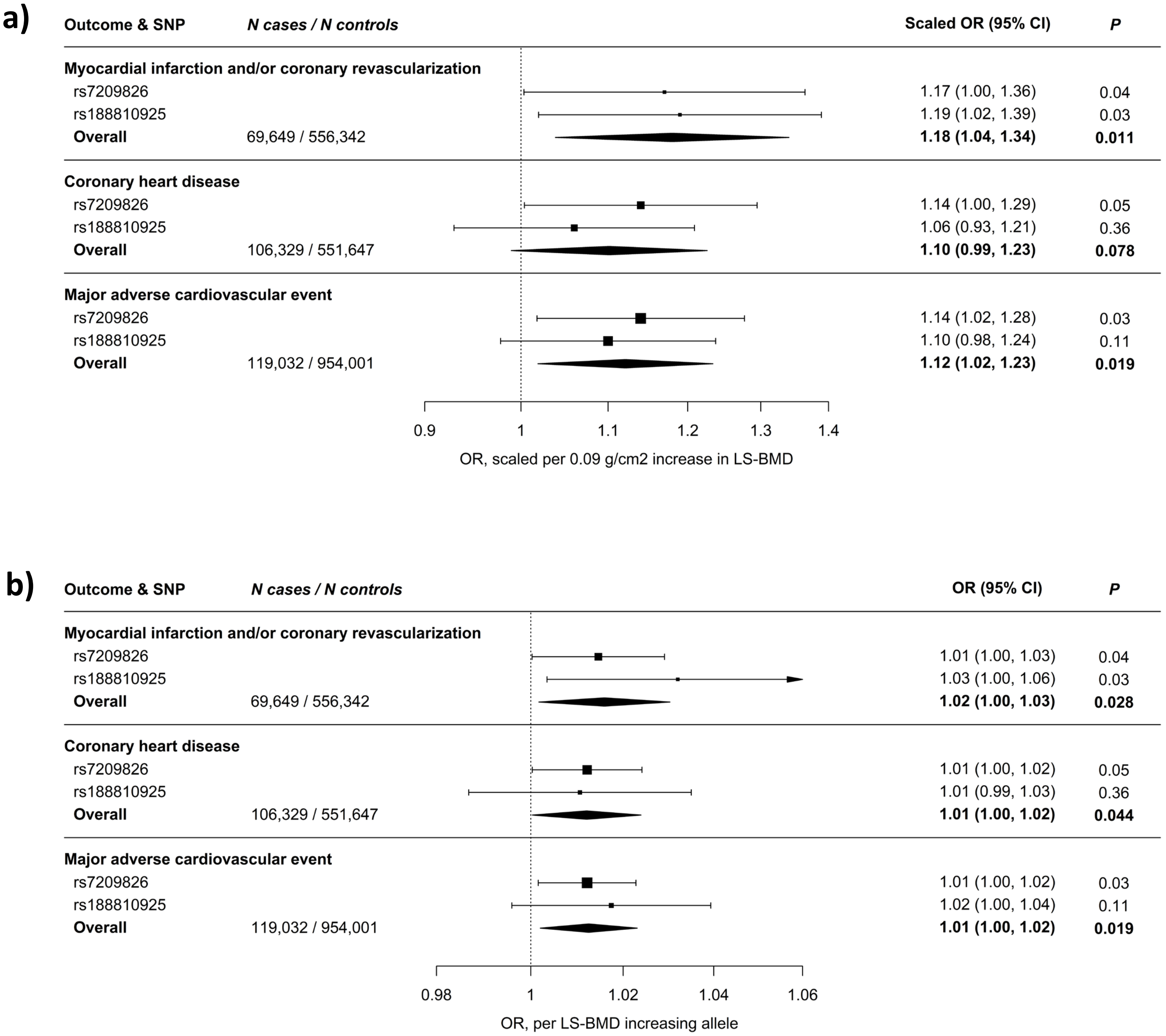


**Figure S10: Meta-analysis of BMD-increasing *SOST* variants with risk of MI and/or coronary revascularization, CHD, and MACE.** a) Association estimates are scaled to match the effect of romosozumab (210 mg monthly for 12 months) on LS-BMD (0.09 g/cm$^2$) and aligned to the BMD-increasing alleles. b) Association estimates on the original allelic scale. MI and/or coronary revascularization includes codes pertaining to MI, coronary artery bypass graft surgery, and/or percutaneous transluminal coronary angioplasty; up to 69,649 cases were analysed. CHD includes all codes pertaining to MI and/or coronary revascularization, as well as codes for angina and chronic stable ischemic heart disease; up to 106,329 cases were analysed. MACE includes codes pertaining to a composite of MI and/or coronary revascularization, stroke, or death from either; up to 119,032 cases were analysed. Boxes represent point estimates of effects. Lines represent 95% confidence intervals.
